# Diagnostic and prognostic value of ECG-predicted hypertension-mediated left ventricular hypertrophy using machine learning

**DOI:** 10.1101/2024.04.22.24306204

**Authors:** Hafiz Naderi, Julia Ramírez, Stefan van Duijvenboden, Esmeralda Ruiz Pujadas, Nay Aung, Lin Wang, Bishwas Chamling, Marcus Dörr, Marcello R P Markus, C. Anwar A Chahal, Karim Lekadir, Steffen E Petersen, Patricia B Munroe

## Abstract

**Background:** Four hypertension-mediated left ventricular hypertrophy (LVH) phenotypes have been reported using cardiac magnetic resonance (CMR): normal LV, LV remodeling, eccentric and concentric LVH, with varying prognostic implications. The electrocardiogram (ECG) is routinely used to detect LVH, however its capacity to differentiate between LVH phenotypes is unknown. This study aimed to classify hypertension-mediated LVH from the ECG using machine learning (ML) and test for associations of ECG-predicted phenotypes with incident cardiovascular outcomes.

**Methods:** ECG biomarkers were extracted from the 12-lead ECG of 20,439 hypertensives in UK Biobank (UKB). Classification models integrating ECG and clinical variables were built using logistic regression, support vector machine (SVM) and random forest. The models were trained in 80% of the participants, and the remaining 20% formed the test set. External validation was sought in 877 hypertensives from the Study of Health in Pomerania (SHIP). In the UKB test set, we tested for associations between ECG-predicted LVH phenotypes and incident major adverse cardiovascular events (MACE) and heart failure.

**Results:** Among UKB participants 19,408 had normal LV, 758 LV remodeling, 181 eccentric and 92 concentric LVH. Classification performance of the three models was comparable, with SVM having a slightly superior performance (accuracy 0.79, sensitivity 0.59, specificity 0.87, AUC 0.69) and similar results observed in SHIP. There was superior prediction of eccentric LVH in both cohorts. In the UKB test set, ECG-predicted eccentric LVH was associated with heart failure (HR 3.42, CI 1.06-9.86).

**Conclusions:** ECG-based ML classifiers represent a potentially accessible screening strategy for the early detection of hypertension-mediated LVH phenotypes.

**Graphical abstract:** 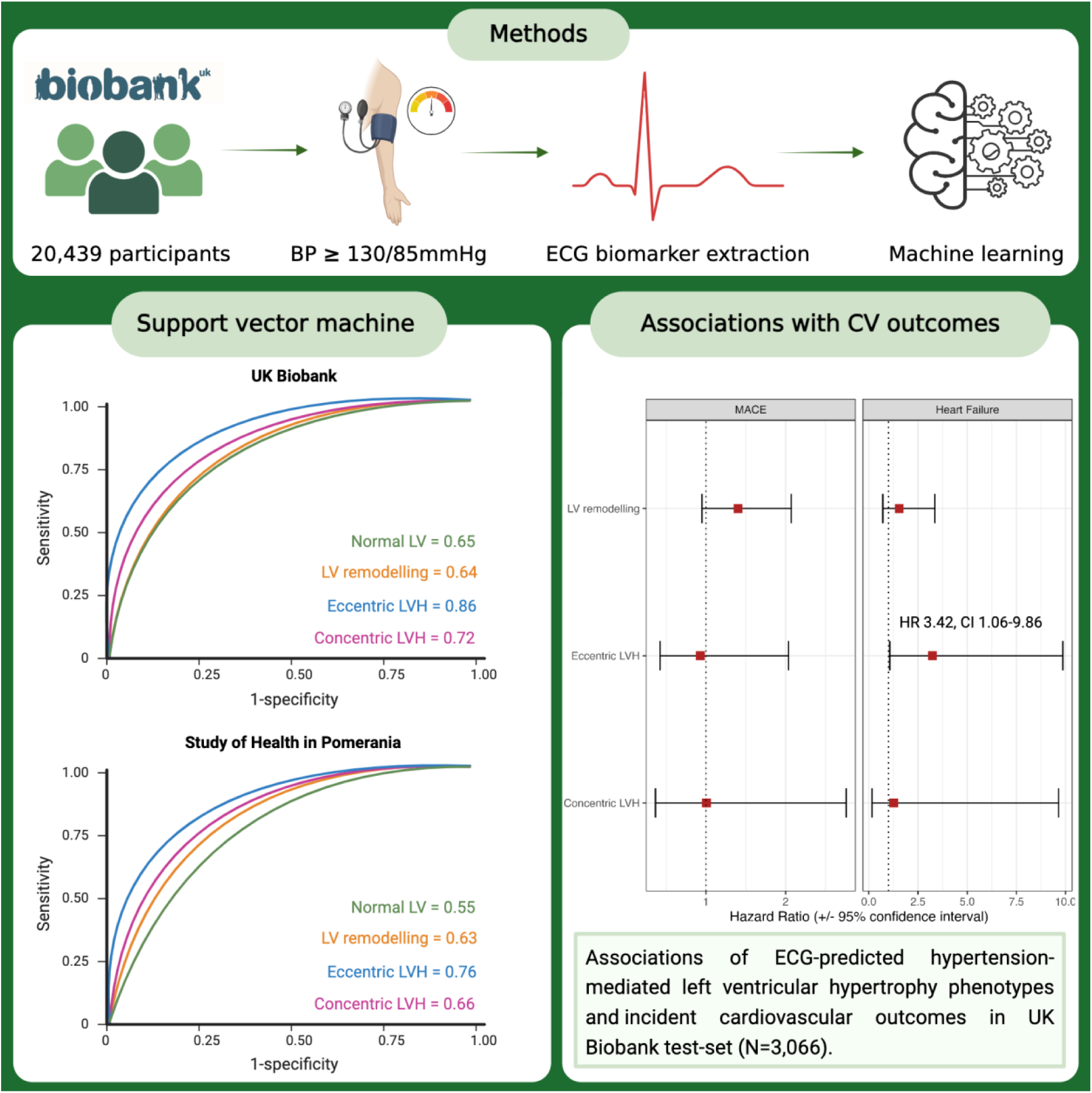

## Introduction

Hypertension is the most common cause of left ventricular hypertrophy (LVH), and both are strong predictors of cardiovascular (CV) morbidity and mortality.^1,2^ The diagnosis of hypertension-mediated LVH has relied on cardiac imaging, such as echocardiography and cardiac magnetic resonance (CMR).^3,4^ Using CMR imaging, four distinct hypertension-mediated LVH phenotypes have been reported: normal left ventricle (LV), LV remodeling, eccentric LVH and concentric LVH.^5^ The spectrum of LVH phenotypes has been shown to have varying prognostic implications, with worse CV outcomes reported in eccentric and concentric LVH.^6,7^ Due to the global burden of hypertension, a cost-effective approach in detecting LVH phenotypes is required to meet clinical demand. Before the advent of CV imaging, the electrocardiogram (ECG) had been used clinically to screen for LVH in hypertension.^8–10^ However, its capacity to detect the four CMR-defined LVH phenotypes is unknown.

Hypertension clinical guidelines endorse using the 12-lead ECG in individuals to screen for LVH.^3,4,11^ The ECG is a readily available and low-cost first line diagnostic tool performed on most patients during an acute care visit and follow-up of chronic CV conditions. In recent years, the transition to digitised ECG in electronic healthcare records has paved opportunities for ECG-based diagnostic and prognostic predictions. Moreover, the use of wearable technology and smartphones have increased its accessibility. Early detection of hypertension-mediated LVH can enable regular healthcare follow-up, rigorous CV risk management and timely initiation of effective blood pressure (BP)-reducing therapies. However, accurate reporting of the ECG is challenging for clinicians, and any improvement in automated analysis could ensure timely diagnosis and treatment of hypertensive patients with LVH.^12–14^ A machine learning (ML) tool to detect hypertension-mediated LVH phenotypes could reduce the number of unnecessary CMR scans, allowing them to be used more efficiently, thus reducing waiting times. This is also of clinical significance as the ECG features derived from classifying hypertension-mediated LVH phenotypes may be used as surrogate markers to predict clinical outcomes in hypertensives.

This study uses ML techniques to explore the diagnostic and prognostic value of ECG-predicted hypertension-mediated LVH phenotypes. We hypothesized that a selection of ECG biomarkers and clinical variables could classify hypertension-mediated LVH phenotypes defined by CMR imaging and that these ECG-predicted LVH phenotypes would be associated with incident CV outcomes.

## Methods

### UK Biobank sample selection

The UK Biobank (UKB) is a large prospective population study where demographics, medication history, electronic health records, biomarkers and genomics were collected in half a million participants aged 40-69 years when recruited between 2006 and 2010 from across the United Kingdom. The UKB imaging study was launched in 2015 with the aim of scanning 20% of the original cohort, that is 100,000 participants.^15^ The details of the UKB CMR protocol have been described elsewhere.^16^

A total of 44,817 participants had completed the UKB imaging study at the time of analysis. Of these, 37,651 participants had both ECG and CMR data available. Hypertensive participants (N=23,042) were identified according to the ‘high normal’ BP grade of ≥130/85mmHg based on the 2018 European Society of Cardiology/European Society of Hypertension (ESC/ESH) guideline.^3^ In UKB, BP readings from the imaging visit were analyzed as these were taken on the same day as the CMR study and 12-lead resting ECG. Each participant had two manual BP readings using a validated automated BP monitor or a manual sphygmomanometer. After calculating the average BP values, we adjusted for medication use by adding 15 and 10 mmHg to SBP and DBP, respectively, for participants reported to be taking BP-lowering medication.^17^ We further defined hypertension by selecting relevant data fields from the UKB data showcase, including hypertension self-reported by participants, formal diagnosis from primary care physician and BP medication use. Participants with other causes of LVH (N=2,603) were excluded by reviewing exome sequence data for genes implicated in hypertrophic cardiomyopathy.^18^ The remaining hypertensive participants (N=20,439) were categorized into the four hypertension-mediated LVH phenotypes using a mass-to-volume ratio. Indexing for body surface area was performed using the Mosteller formula.^19^ The data fields selected from UKB are shown in Supplementary Table 1. Figure 1 illustrates the UKB sample selection process.

**Figure 1.**
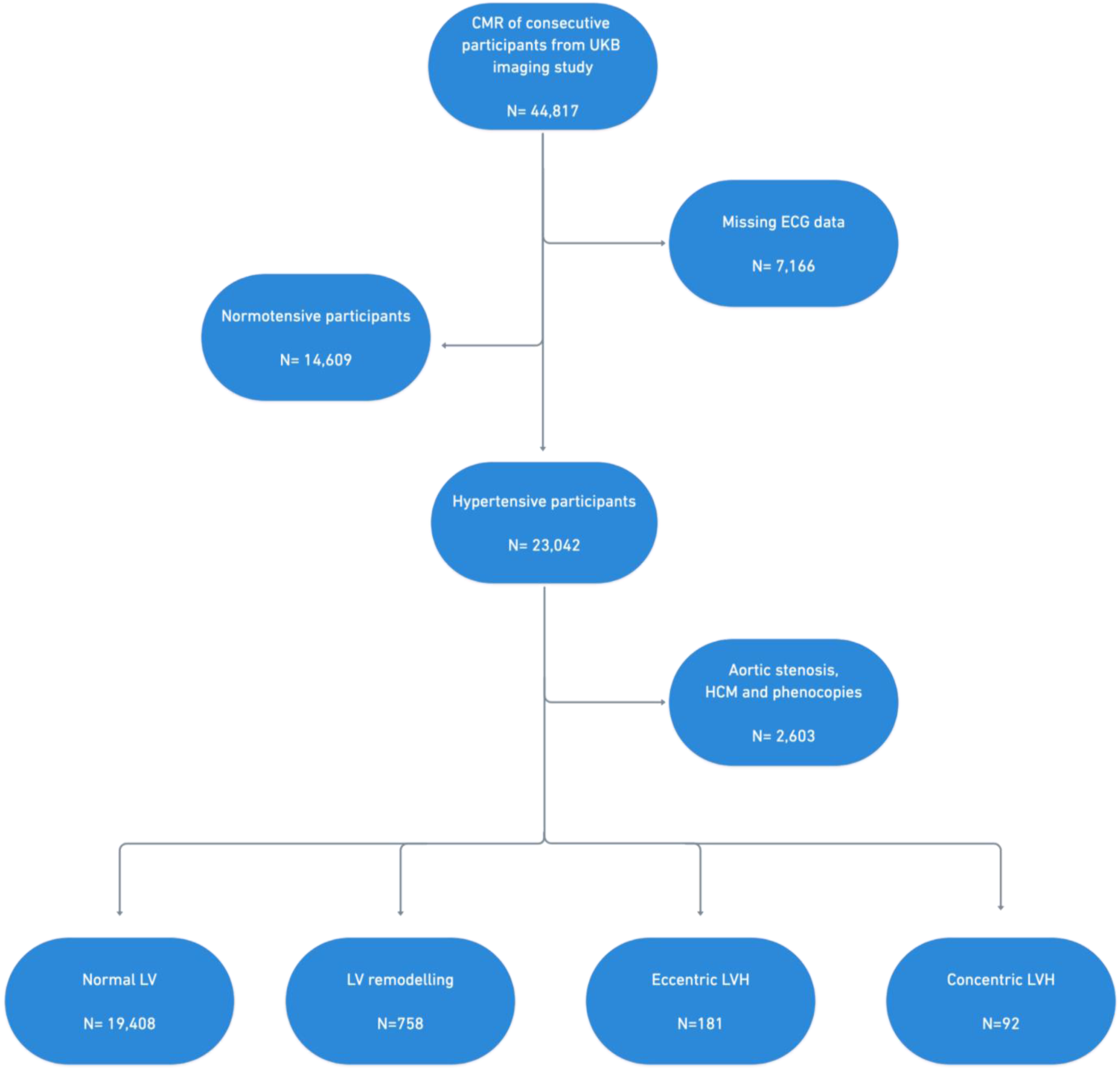
Flow diagram illustrating the steps involved in UK Biobank participant selection. *CMR: cardiac magnetic resonance imaging; HCM: hypertrophic cardiomyopathy; LV: left ventricle; LVH: left ventricular hypertrophy*.

### ECG biomarker extraction

Full 15-second 12-lead ECG signals of each of the 20,739 participants were analyzed semi-automatically using a custom algorithm written in MATLAB (version 2021a, Mathworks Inc.) to derive 23 biomarkers (Supplementary Table 2) with a known physiological association with LVH.^20^ Only the independent ECG leads (I, II, V1-6) were analyzed. Butterworth filter (1-45Hz) was applied to attenuate baseline wander and high-frequency noise. ECG biomarkers from each independent lead were treated as individual features. In addition, global ECG features were calculated as the median value across the independent leads. The applied algorithm for extracting ECG biomarkers is detailed elsewhere.^21^

### Ascertainment of clinical variables

In addition to ECG biomarkers, we also included clinical variables known to be associated with LVH (**Table 1**) in the classification model. Each clinical variable was defined by either a self-reported questionnaire or biochemistry results. Participants with serum total cholesterol of ≥5mmol/L and Hemoglobin A1c (HbA1c) ≥48 mmol/mol at the baseline visit were considered to have hypercholesterolemia and diabetes mellitus, respectively. We corrected total and non-HDL cholesterol values for participants on cholesterol-lowering medication by dividing the total cholesterol by 0.73 and non-HDL cholesterol by 0.66.^22^ The presence of tobacco use was ascertained using self-reported questionnaires, with smoking status classified categorically as current, previous or never. Alcohol consumption was classified as current or never.

**Table 1.**
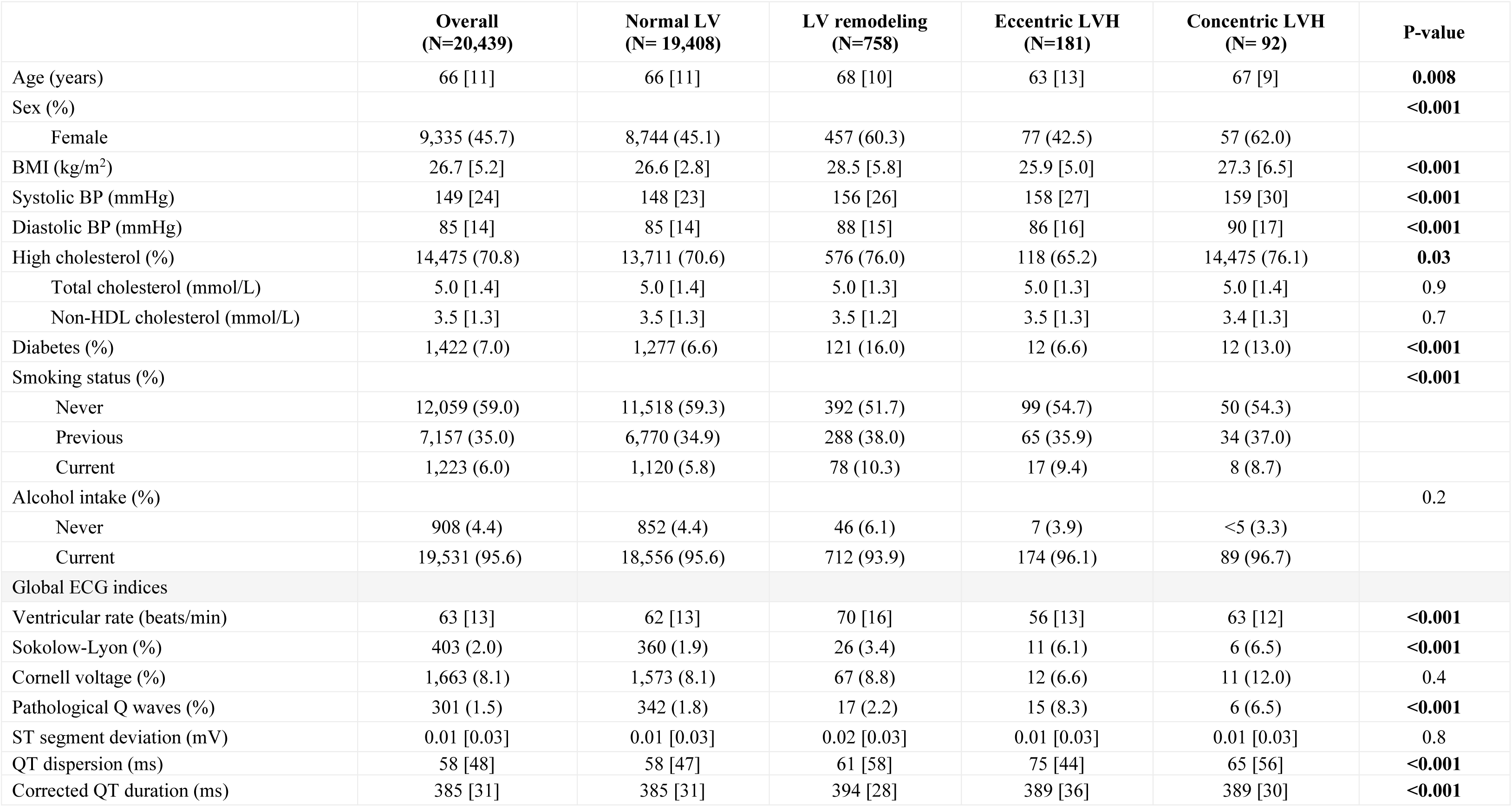
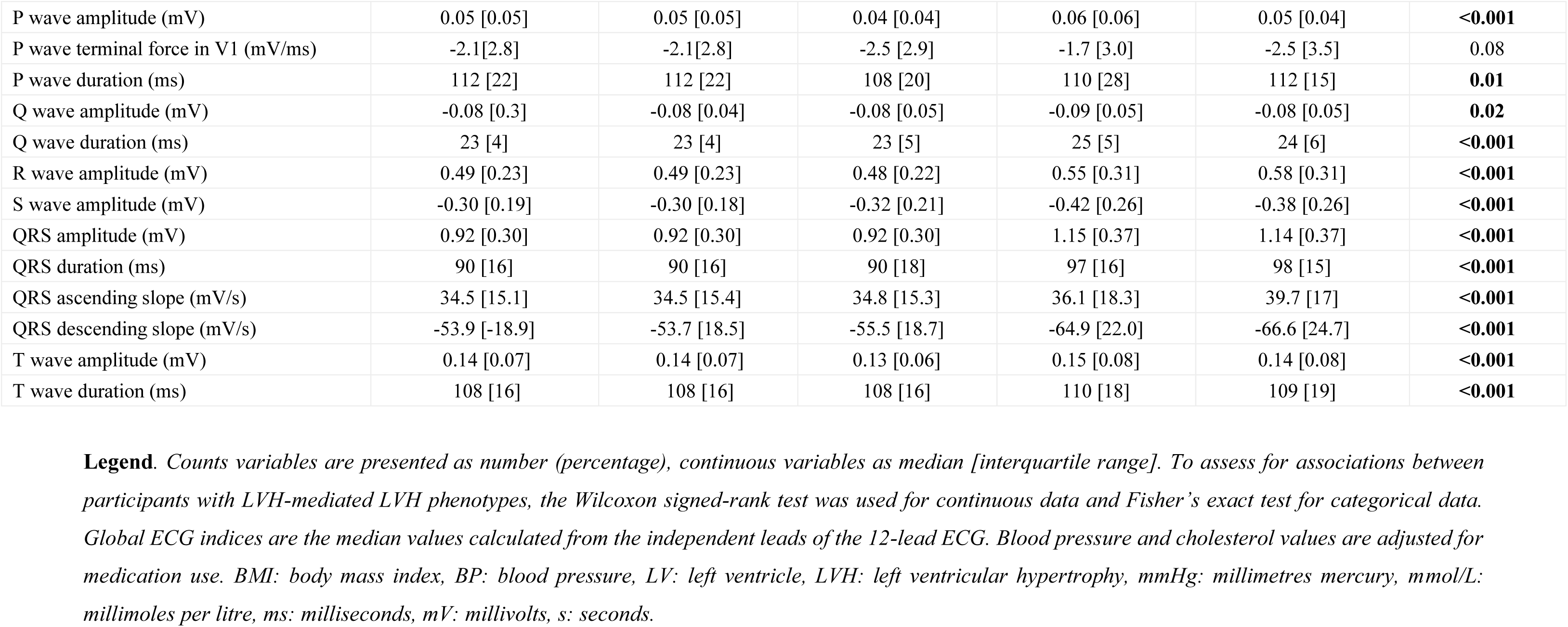
Baseline characteristics of UK Biobank participants.

|  | **Overall**<br>**(N=20,439)** | **Normal LV**<br>**(N= 19,408)** | **LV remodeling**<br>**(N=758)** | **Eccentric LVH**<br>**(N=181)** | **Concentric LVH**<br>**(N= 92)** | **P-value** |
| --- | --- | --- | --- | --- | --- | --- |
| Age (years) | 66 [11] | 66 [11] | 68 [10] | 63 [13] | 67 [9] | **0.008** |
| Sex (%) |  |  |  |  |  | **<0.001** |
| Female | 9,335 (45.7) | 8,744 (45.1) | 457 (60.3) | 77 (42.5) | 57 (62.0) |  |
| BMI (kg/m<sup>2</sup>) | 26.7 [5.2] | 26.6 [2.8] | 28.5 [5.8] | 25.9 [5.0] | 27.3 [6.5] | **<0.001** |
| Systolic BP (mmHg) | 149 [24] | 148 [23] | 156 [26] | 158 [27] | 159 [30] | **<0.001** |
| Diastolic BP (mmHg) | 85 [14] | 85 [14] | 88 [15] | 86 [16] | 90 [17] | **<0.001** |
| High cholesterol (%) | 14,475 (70.8) | 13,711 (70.6) | 576 (76.0) | 118 (65.2) | 14,475 (76.1) | **0.03** |
| Total cholesterol (mmol/L) | 5.0 [1.4] | 5.0 [1.4] | 5.0 [1.3] | 5.0 [1.3] | 5.0 [1.4] | 0.9 |
| Non-HDL cholesterol (mmol/L) | 3.5 [1.3] | 3.5 [1.3] | 3.5 [1.2] | 3.5 [1.3] | 3.4 [1.3] | 0.7 |
| Diabetes (%) | 1,422 (7.0) | 1,277 (6.6) | 121 (16.0) | 12 (6.6) | 12 (13.0) | **<0.001** |
| Smoking status (%) |  |  |  |  |  | **<0.001** |
| Never | 12,059 (59.0) | 11,518 (59.3) | 392 (51.7) | 99 (54.7) | 50 (54.3) |  |
| Previous | 7,157 (35.0) | 6,770 (34.9) | 288 (38.0) | 65 (35.9) | 34 (37.0) |  |
| Current | 1,223 (6.0) | 1,120 (5.8) | 78 (10.3) | 17 (9.4) | 8 (8.7) |  |
| Alcohol intake (%) |  |  |  |  |  | 0.2 |
| Never | 908 (4.4) | 852 (4.4) | 46 (6.1) | 7 (3.9) | <5 (3.3) |  |
| Current | 19,531 (95.6) | 18,556 (95.6) | 712 (93.9) | 174 (96.1) | 89 (96.7) |  |
| Global ECG indices |  |  |  |  |  |  |
| Ventricular rate (beats/min) | 63 [13] | 62 [13] | 70 [16] | 56 [13] | 63 [12] | **<0.001** |
| Sokolow-Lyon (%) | 403 (2.0) | 360 (1.9) | 26 (3.4) | 11 (6.1) | 6 (6.5) | **<0.001** |
| Cornell voltage (%) | 1,663 (8.1) | 1,573 (8.1) | 67 (8.8) | 12 (6.6) | 11 (12.0) | 0.4 |
| Pathological Q waves (%) | 301 (1.5) | 342 (1.8) | 17 (2.2) | 15 (8.3) | 6 (6.5) | **<0.001** |
| ST segment deviation (mV) | 0.01 [0.03] | 0.01 [0.03] | 0.02 [0.03] | 0.01 [0.03] | 0.01 [0.03] | 0.8 |
| QT dispersion (ms) | 58 [48] | 58 [47] | 61 [58] | 75 [44] | 65 [56] | **<0.001** |
| Corrected QT duration (ms) | 385 [31] | 385 [31] | 394 [28] | 389 [36] | 389 [30] | **<0.001** |
| P wave amplitude (mV) | 0.05 [0.05] | 0.05 [0.05] | 0.04 [0.04] | 0.06 [0.06] | 0.05 [0.04] | **<0.001** |
| P wave terminal force in V1 (mV/ms) | -2.1[2.8] | -2.1[2.8] | -2.5 [2.9] | -1.7 [3.0] | -2.5 [3.5] | 0.08 |
| P wave duration (ms) | 112 [22] | 112 [22] | 108 [20] | 110 [28] | 112 [15] | **0.01** |
| Q wave amplitude (mV) | -0.08 [0.3] | -0.08 [0.04] | -0.08 [0.05] | -0.09 [0.05] | -0.08 [0.05] | **0.02** |
| Q wave duration (ms) | 23 [4] | 23 [4] | 23 [5] | 25 [5] | 24 [6] | **<0.001** |
| R wave amplitude (mV) | 0.49 [0.23] | 0.49 [0.23] | 0.48 [0.22] | 0.55 [0.31] | 0.58 [0.31] | **<0.001** |
| S wave amplitude (mV) | -0.30 [0.19] | -0.30 [0.18] | -0.32 [0.21] | -0.42 [0.26] | -0.38 [0.26] | **<0.001** |
| QRS amplitude (mV) | 0.92 [0.30] | 0.92 [0.30] | 0.92 [0.30] | 1.15 [0.37] | 1.14 [0.37] | **<0.001** |
| QRS duration (ms) | 90 [16] | 90 [16] | 90 [18] | 97 [16] | 98 [15] | **<0.001** |
| QRS ascending slope (mV/s) | 34.5 [15.1] | 34.5 [15.4] | 34.8 [15.3] | 36.1 [18.3] | 39.7 [17] | **<0.001** |
| QRS descending slope (mV/s) | -53.9 [-18.9] | -53.7 [18.5] | -55.5 [18.7] | -64.9 [22.0] | -66.6 [24.7] | **<0.001** |
| T wave amplitude (mV) | 0.14 [0.07] | 0.14 [0.07] | 0.13 [0.06] | 0.15 [0.08] | 0.14 [0.08] | **<0.001** |
| T wave duration (ms) | 108 [16] | 108 [16] | 108 [16] | 110 [18] | 109 [19] | **<0.001** |

**Legend.** Counts variables are presented as number (percentage), continuous variables as median [interquartile range]. To assess for associations between participants with LVH-mediated LVH phenotypes, the Wilcoxon signed-rank test was used for continuous data and Fisher's exact test for categorical data. Global ECG indices are the median values calculated from the independent leads of the 12-lead ECG. Blood pressure and cholesterol values are adjusted for medication use. BMI: body mass index, BP: blood pressure, LV: left ventricle, LVH: left ventricular hypertrophy, mmHg: millimetres mercury, mmol/L: millimoles per litre, ms: milliseconds, mV: millivolts, s: seconds.

### Supervised machine learning techniques

Three supervised ML algorithms were evaluated for classification: logistic regression, support vector machine (SVM) and random forest (RF). The algorithms were implemented in MATLAB, and the fit multiclass models for SVMs or other classifiers (fitcecoc) function was used to build the logistic regression and SVM classifiers.^23^ The fit ensemble of learners for classification (fitcensemble) was used to build the RF classifier.^24^ In our experiments, the dataset was split into a training set (80%) for learning and a test set (20%) for performance evaluation. We applied 10-fold cross-validation to the training set. The metrics we used to assess classifier performance in the test set included: accuracy, sensitivity, specificity, precision, F1 score and area under the receiver operator curve (AUC). Further details of the models are described previously.^21^

### External validation in Study of Health in Pomerania

The Study of Health in Pomerania (SHIP) is a population-based study investigating common risk factors and subclinical diseases from a random cluster sample drawn from the population of West Pomerania in Northeast of Germany.^25,26^ SHIP consists of two independent cohorts: SHIP-START (recruited between 1997 to 2001) and SHIP-TREND (recruited between 2008 to 2012). Study participants for both cohorts were sampled from the general adult population aged 20-79 in West Pomerania. This study used the second follow-up of SHIP-START (SHIP-START-2) and baseline SHIP-TREND-0 as these were the studies with both 12-lead ECG and CMR data. The populations comprised 2,333 participants for SHIP-START-2 and 4,420 participants for SHIP-TREND-0.^26^ A total of 1,474 participants from SHIP had both CMR and ECG data. The same definition of hypertension was applied based on BP readings, medication use and diagnosis, yielding a total of 877 hypertensives in SHIP. The same ECG biomarkers and clinical features as per UKB analysis were extracted. For classification, the whole SHIP cohort was treated as a test set. Therefore, down-sampling was not applied. The best-performing ML model (SVM) was taken forward for external validation in SHIP.

### Associations with cardiovascular outcomes in UK Biobank

Longitudinal data on clinical outcomes of UKB participants is recorded using linkage to Hospital Episode Statistics (HES) and the UK death register.^27^ All UKB participants consented to be followed up. In this study, the primary endpoint was major adverse cardiovascular events (MACE), defined as either hospitalization or death due to fatal/non-fatal myocardial infarction, stroke or ventricular arrhythmias. Cases were identified using relevant International Classification of Disease, 9^th^ or 10^th^ Revision (ICD-9, ICD-10), or Office of Population Censuses and Surveys version 4 (OPCS 4) Classification of Interventions and Procedures codes in the health-related records or death register (Supplementary Table 3). An additional analysis was performed testing for association with heart failure separately. These clinical outcomes were selected due to their association with hypertension from the literature and clinical expertise. The follow-up period was determined by the first appearance of ICD-9, ICD-10 or OPSC4 codes in either health record or death register data since the UKB imaging visit. Participants with prevalent events at the time of UKB enrolment were excluded from the survival analyses. Participants who did not experience an event were censored at death or the end of the follow-up period (30 November 2022).

### Statistical analyses

Statistical analyses were performed using R version 4.0.3 and RStudio Version 1.3.1093.^28^ After excluding missing or extreme ECG values (outside the range defined by the quartiles +/- 1.5 x interquartile range) the Classification And REgression Training (CARET) package in R was used for correlation analysis, and highly correlated ECG biomarkers were omitted (correlation coefficient threshold of +/- 0.9).^29^ ECG biomarkers with less than 10% of missing data were imputed using the Multivariate Imputation by Chained Equations (MICE) package in R.^30^ In order to address the imbalance in the dataset, down-sampling was applied using the CARET package in the training set to match the proportion of participants in the minority LVH group. A chi-squared test was used to rank the features in terms of feature importance score.

Descriptive statistics are presented as median (interquartile range) for continuous variables or frequency (percentage) for categorical variables. The distribution of continuous data was assessed by visual inspection of the histograms and confirmed by the Shapiro-Wilk test. Baseline clinical and ECG characteristics of the hypertension-mediated LVH phenotypes were statistically compared with the normal LV group. To assess for associations, the ANOVA test was used for continuous data and the chi-squared test for categorical data. For all analyses, a two-tailed p-value <0.05 was deemed statistically significant.

Associations between the ECG-predicted phenotypes and clinical outcomes were performed in the UKB test set (N = 3,066) using multivariable-adjusted Cox proportional hazard regression, setting normal LV as the reference group. For each clinical outcome the model was adjusted for age, sex and body mass index. Hazard ratios (HR) were reported with 95% confidence intervals (CI) to derive risk for each LVH phenotype compared to the normal LV group.

## Results

### Study population

The clinical and ECG characteristics of the UKB participants stratified by hypertension-mediated LVH phenotypes are presented in Table 1. Among the 20,439 hypertensive participants in UKB, 19,408 (95.0%)had normal LV, 758 (3.7%) had LV remodeling, 181 (0.9%) eccentric LVH and 92 (0.5%) concentric LVH. Overall, the cohort had an average age of 66 years, and 46% were female. In the total hypertensive cohort, the frequency of participants with LVH criteria for Sokolow-Lyon and Cornell voltage on the ECG was 2% and 8%, respectively.

Table 2 shows the baseline characteristics of the SHIP validation cohort. In SHIP there were 877 participants with hypertension, of which 704 (80.3%) had normal LV, 134 (15.3%) LV remodeling, 12 (1.4%) participants with eccentric LVH and 27 (3.1%) with concentric LVH. The average age was 56 years, and 39% were female. Overall, the frequency of participants with LVH criteria for Sokolow-Lyon and Cornell voltage on the ECG was 6% and 9%, respectively.

**Table 2.** Baseline characteristics of particiants in the Study of Health in Pomerania.

|  | **Overall**<br>**(N=877)** | **Normal LV**<br>**(N= 704)** | **LV remodeling**<br>**(N=134)** | **Eccentric LVH**<br>**(N=12)** | **Concentric LVH**<br>**(N= 27)** | **P-value** |
| --- | --- | --- | --- | --- | --- | --- |
| Age (years) | 56 [19] | 55 [20] | 61 [16] | 49 [27] | 59 [15] | **0.02** |
| Sex (%) |  |  |  |  |  | **0.04** |
| Female | 344 (39.2) | 261 (37.1) | 67 (50.0) | 6 (50.0) | 10 (37.0) |  |
| BMI (kg/m<sup>2</sup>) | 28.0 [5.4] | 27.7 [5.4] | 29.5 [5.1] | 25.1 [4.3] | 30.0 [5.8] | **0.007** |
| Systolic BP (mmHg) | 135 [18] | 135 [18] | 135 [22] | 132 [22] | 145 [20] | **<0.001** |
| Diastolic BP (mmHg) | 82 [13] | 82 [12] | 81 [15] | 75 [17] | 83 [12] | 0.3 |
| High cholesterol (%) | 693 (79.0) | 542 (77.0) | 121 (90.3) | 8 (66.7) | 22 (81.5) | **0.004** |
| Total cholesterol (mmol/L) | 5.8 [1.4] | 5.8 [1.3] | 6.1 [1.4] | 5.4 [1.6] | 6.0 [1.5] | **0.01** |
| Non-HDL cholesterol (mmol/L) | 4.4 [1.3] | 4.3 [1.4] | 4.6 [1.2] | 4.0 [2.1] | 4.7 [1.3] | **0.01** |
| Diabetes (%) | 772 (88.0) | 618 (87.8) | 120 (89.6) | 9 (75.0) | 25 (92.6) | 0.4 |
| Smoking status (%) |  |  |  |  |  | **0.008** |
| Previous | 728 (83.0) | 597 (84.8) | 105 (78.4) | 9 (75.0) | 17 (63.0) |  |
| Current | 149 (17.0) | 107 (15.2) | 29 (21.6) | <5 (25.0) | 10 (37.0) |  |
| Alcohol intake (%) |  |  |  |  |  | 0.2 |
| Never | 53 (6.0) | 41 (5.8) | 10 (7.5) | <5 (16.7) | 0 (0) |  |
| Current | 824 (94.0) | 663 (94.2) | 124 (92.5) | 10 (83.3) | 27 (100.0) |  |
| Global ECG indices |  |  |  |  |  |  |
| Ventricular rate (beats/min) | 64 [14] | 63 [14] | 68 [13] | 62 [10] | 65 [16] | **0.04** |
| Sokolow-Lyon (%) | 50 (5.7) | 37 (5.3) | 6 (4.5) | <5 (25.0) | <5 (14.8) | **0.004** |
| Cornell voltage (%) | 75 (8.6) | 62 (8.8) | 10 (7.5) | <5 (8.3) | <5 (7.4) | 0.9 |
| ST segment deviation (mV) | 0.03 [0.03] | 0.03 [0.03] | 0.03 [0.03] | 0.03 [0.05] | 0.03 [0.03] | 0.7 |
| QT dispersion (ms) | 47 [45] | 44 [45] | 45 [51] | 72 [22] | 60 [39] | **0.04** |
| Corrected QT duration (ms) | 384 [27] | 384 [29] | 387 [26] | 378 [36] | 387 [26] | 0.2 |
| P wave amplitude (mV) | 0.02 [0.03] | 0.02 [0.03] | 0.02 [0.03] | 0.01 [0.03] | 0.03 [0.01] | 0.7 |
| P wave terminal force in V1 (mV/ms) | -2.8 [2.3] | -2.8 [2.3] | -3.2 [2.2] | -1.8 [1.6] | -2.5 [2.1] | 0.5 |
| P wave duration (ms) | 86 [20] | 86 [18] | 86 [20] | 79 [12] | 86 [23] | 0.8 |
| Q wave amplitude (mV) | -0.10 [0.05] | -0.10 [0.05] | -0.09 [0.04] | -0.10 [0.06] | -0.12 [0.05] | 0.3 |
| Q wave duration (ms) | 24 [4] | 24 [4] | 24 [4] | 24 [3] | 23 [4] | 0.3 |
| R wave amplitude (mV) | 0.55 [0.24] | 0.55 [0.23] | 0.53 [0.26] | 0.65 [0.18] | 0.63 [0.38] | 0.4 |
| S wave amplitude (mV) | -0.24 [0.16] | -0.23 [0.16] | -0.25 [0.18] | -0.24 [0.13] | -0.28 [0.18] | 0.1 |
| QRS amplitude (mV) | 0.98 [0.35] | 0.98 [0.33] | 0.96 [0.34] | 1.22 [0.27] | 1.20 [0.40] | 0.7 |
| QRS duration (ms) | 90 [10] | 90 [10] | 89 [9] | 90 [10] | 92 [9] | 0.1 |
| QRS ascending slope (mV/s) | 37.9 [14.9] | 37.6 [14.3] | 38.1 [17.3] | 41.1 [9.0] | 44.0 [25.9] | 0.4 |
| QRS descending slope (mV/s) | -55.1 [19.6] | -54.7 [18.8] | -54.8 [21.5] | -67.1 [21.9] | -62.0 [26.6] | 0.1 |
| T wave amplitude (mV) | 0.16 [0.08] | 0.16 [0.08] | 0.15 [0.07] | 0.16 [0.13] | 0.14 [0.08] | 0.6 |
| T wave duration (ms) | 112 [18] | 112 [18] | 108 [16] | 120 [12] | 110 [21] | 0.4 |

**Legend.** Counts variables are presented as number (percentage), continuous variables as median [interquartile range]. To assess for associations between participants with LVH-mediated LVH phenotypes, the Wilcoxon signed-rank test was used for continuous data and Fisher's exact test for categorical data. Global ECG indices are the median values calculated from the independent leads of the 12-lead ECG. Blood pressure and cholesterol values are adjusted for medication use. BMI: body mass index, BP: blood pressure, LV: left ventricle, LVH: left ventricular hypertrophy, mmHg: millimetres mercury, mmol/L: millimoles per litre, ms: milliseconds, mV: millivolts, s: seconds.

### Machine learning model performance in UK Biobank

Supplementary Figure 1 shows the ranking of the top 40 features across all ML models using chi-squared feature selection. The top clinical features were sex and age, and the highest-ranking ECG predictors of LVH were ventricular rate and QRS amplitude in V4.

The performance metrics of the supervised ML classifiers (logistic regression, SVM and RF) using both ECG and clinical variables in UKB are shown in Table 3. Classification with each method was comparable in the test set, with SVM showing the most consistent performance with 0.79 accuracy, 0.59 sensitivity, 0.87 specificity, 0.78 precision, 0.67 F1 score and AUC 0.69. Using SVM, the classification of eccentric LVH (AUC 0.86) and concentric LVH (0.72) was superior to normal LV (0.65) and LV remodeling (0.64) phenotypes, as shown in Figure 2. The ECG biomarkers enhanced the model performance of the SVM classifier in UKB, compared to clinical variables alone (AUC values of 0.69 and 0.58, respectively), as illustrated in Figure 2.

**Figure 2.**
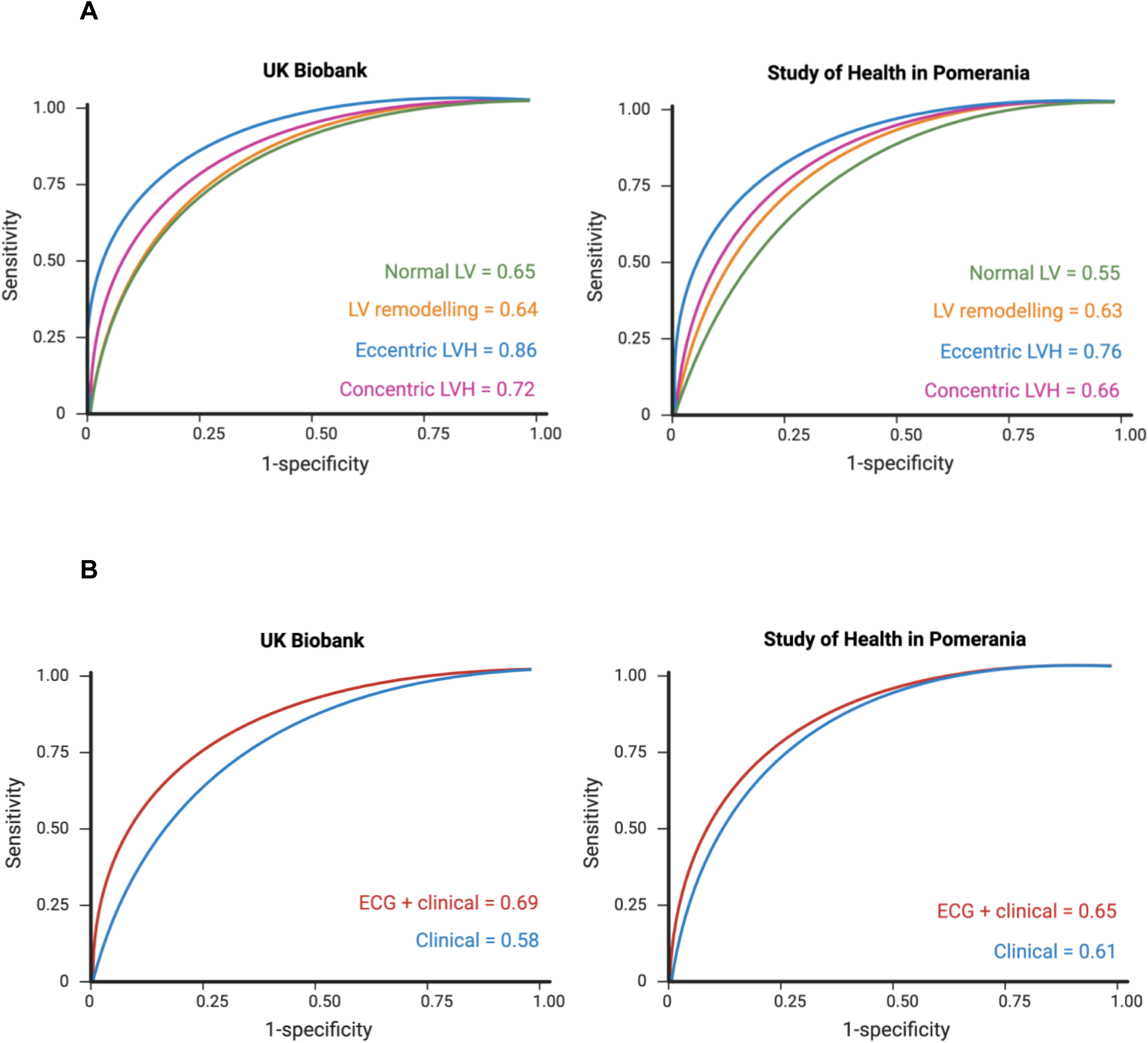
**(A) Classification of hypertension mediated LVH phenotypes in UK Biobank and Study of Health in Pomerania using support vector machine.** **(B) Classification of hypertension mediated LVH phenotypes using ECG and clinical vs clinical data alone in UK Biobank and Study of Health in Pomerania with support vector machine.** *ECG: electrocardiogram; LV: left ventricle; LVH: left ventricular hypertrophy*.

**Table 3.** Performance metrics of supervised machine learning classifiers using ECG and clinical variables in UK Biobank and validation testing in Study of Health in Pomerania.

|  | UK Biobank |  |  | Validation in SHIP |
| --- | --- | --- | --- | --- |
|  | Logistic regression | Support vector machine | Random forest | Support vector machine |
| Accuracy (%) | 0.74 | 0.79 | 0.73 | 0.75 |
| Sensitivity (%) | 0.47 | 0.59 | 0.45 | 0.51 |
| Specificity (%) | 0.83 | 0.87 | 0.82 | 0.85 |
| Precision (%) | 0.68 | 0.78 | 0.64 | 0.63 |
| F1 score (%) | 0.56 | 0.67 | 0.53 | 0.56 |
| AUC | 0.71 | 0.69 | 0.70 | 0.65 |

*AUC: area under the receiver operator curve; LVH: left ventricular hypertrophy; SHIP: Study of Health in Pomerania.*

### External validation in Study of Health in Pomerania

External validation in the SHIP cohort using SVM showed a similar performance to UKB with 0.75 accuracy, 0.51 sensitivity, 0.85 specificity, 0.63 precision, 0.56 F1 score and 0.65 AUC (Table 3). Akin to UKB, the classification of eccentric LVH (AUC 0.76) and concentric LVH (0.66) was superior to normal LV (0.55) and LV remodeling (0.63) phenotypes. The ECG biomarkers also enhanced the model performance of the SVM classifier in the SHIP cohort, compared to clinical variables alone (AUC values of 0.65 and 0.61, respectively, shown in Figure 2).

### Association of ECG-predicted LVH phenotypes with CV outcomes

Figure 3 shows the associations between ECG-predicted hypertension-mediated LVH phenotype using SVM and the clinical outcomes MACE and heart failure in the UKB test set (N = 3,066). There was no statistically significant association with MACE (Supplemental Table 4), however the hazard ratio of heart failure was 3.2 times higher (HR 3.24, CI: 1.06-9.86) in hypertensives with eccentric LVH with normal LV set as the reference group (Supplementary Table 4).

**Figure 3.**
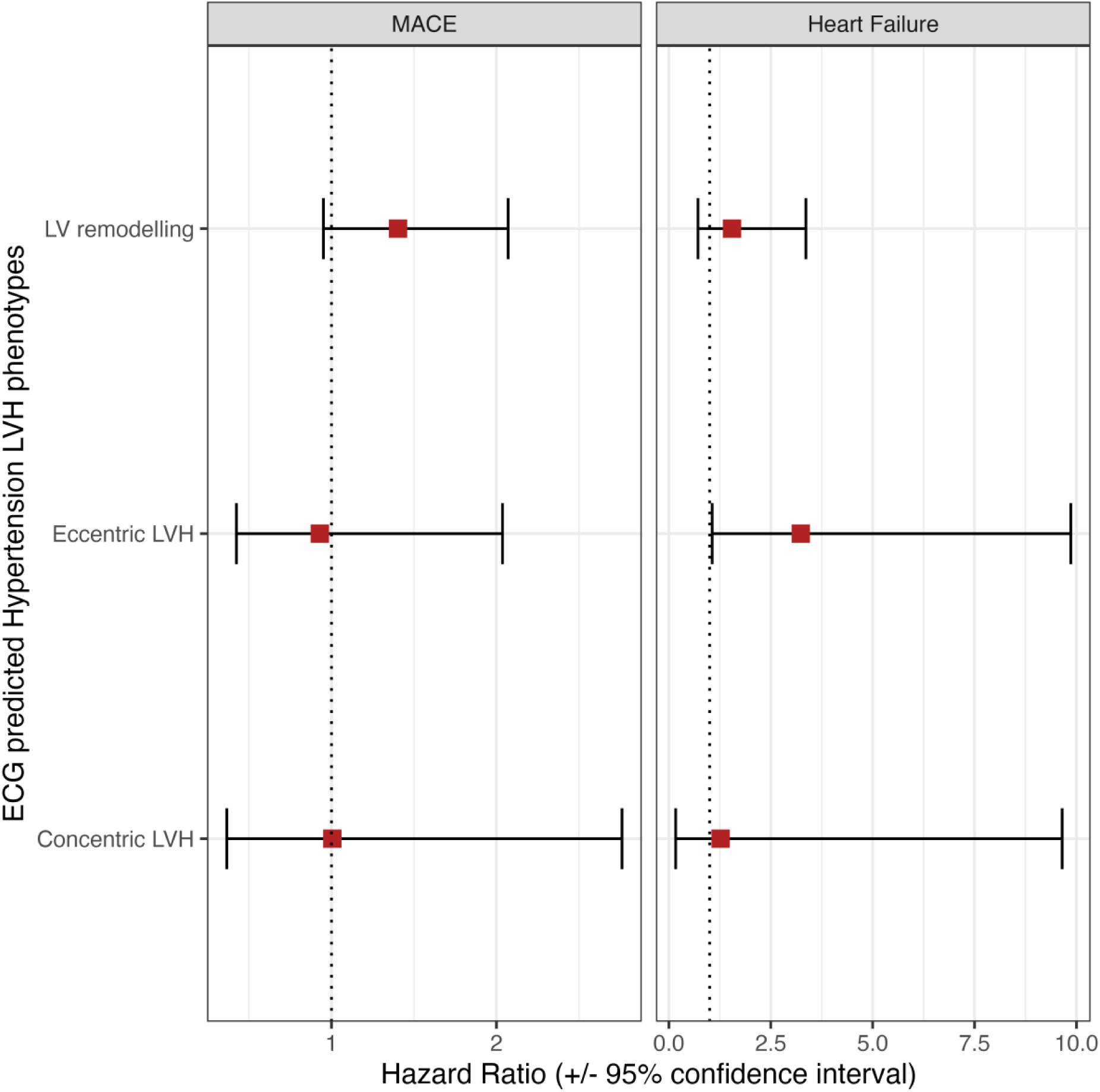
Associations of ECG-predicted hypertension-mediated LVH phenotypes and clinical outcomes. *Results are hazard ratios from Cox hazards proportional regression models. The diseases listed are set as the model outcome (response variable) and hypertension-mediated LV phenotype in the exposure of interest with normal LV as the reference group. The model was adjusted for age, sex and BMI.* *CI: confidence interval; HR: hazard ratio; MACE: major adverse cardiovascular events; LVH: left ventricular hypertrophy*

## Discussion

In UKB, a combination of ECG biomarkers and clinical variables was able to discriminate between hypertension-mediated LVH phenotypes using supervised ML techniques. The ML classifiers had similar performances, with slight superiority using SVM. External validation in the SHIP cohort using SVM demonstrated the robustness of the model with reproducible results. We observed incremental value in using the 12-lead ECG compared to clinical variables alone for hypertension-mediated LVH detection. The classification of eccentric LVH and concentric LVH was superior to normal LV and LV remodeling phenotypes in both UKB and the SHIP cohort. Furthermore, we observed a strong association between the ECG-predicted eccentric LVH group in the UKB test set and heart failure (HR 3.24, CI: 1.06-9.86), indicating there is potential clinical relevance of the model.

This is the first ML study to classify hypertension-mediated LVH phenotypes from the ECG. Ventricular rate and QRS amplitude in the precordial leads were the most influential ECG features in the model. The other top ECG predictors of LVH were measurements relating to the QRS complex, such as QRS duration, QRS descending slope and Sokolow-Lyon criteria, which are derived from amplitude measures of the QRS complex. Change in the QRS complex is a marker of electrical remodeling seen in LVH, which has been postulated to be due to the increase in the muscle mass of the LV mounting the forces of the LV potential. However, the increased QRS voltage is seen only in a minority of LVH cases in both clinical and animal studies, and consequently, voltage criteria suffer from a high number of false negative results and low sensitivity.^31^ In prior work, ECG predictors of LVH have suffered low sensitivity, ranging from 15-30%.^32^ Using a combination of ECG and clinical variables, our sensitivity values were higher, with over 50% using SVM, without compromising specificity (87% using SVM).

The results across all models were comparable, with SVM demonstrating the best overall model performance in classifying hypertension-mediated LVH phenotypes. There is no direct comparative study; however, Beneyto and colleagues (2023) also found SVM to be superior in detecting hypertension as the cause of LVH, compared to the decision tree and RF models.^33^ The authors defined LVH as maximal LV wall thickness greater than 12mm in diastole and found that SVM had the optimal balance between specificity of 86% and sensitivity of 31%. Beneyto *et al* used a combination of clinical, laboratory and ECG features in their models. They identified systolic BP and the number of anti-hypertensive medications among the most significant features for classification. However, as ECG features were not included, this precludes direct comparison with our study.

We validated our findings in an independent cohort, and the results demonstrated robustness of the ML model. Although both cohorts were European, they had differing clinical profiles. Compared to UKB, the SHIP cohort was younger (66 years vs 56 years) and had a higher incidence of hypercholesterolemia (19% vs 79%). The SHIP cohort is noted to be a ‘high-risk’ population compared to the relatively ‘healthy’ cohort of UKB. Despite the differing risk profiles of these cohorts, our model’s comparative performance indicated potential translatability to community populations but also the requirement for further validation experiments.

We speculate that the superior prediction of concentric LVH and eccentric LVH is perhaps due to the distinct geometry of these phenotypes on imaging. A dilated LV characterises eccentric LVH, while concentric LVH is synonymous with a thickened LV wall and small LV cavity size. In contrast, normal LV and LV remodeling are less distinct, hence the inferior classification of these phenotypes using ML.

Following the validation of the model in the SHIP dataset, we were interested in assessing whether the ECG-predicted phenotypes were associated with clinical outcomes. Using the test set in UKB (N=3,066) we observed a significant association between eccentric LVH and heart failure with a 3.2 increased hazard rate of heart failure compared to those with normal LV geometry. Hypertension leads to heart failure through LVH and LV diastolic dysfunction, eventually progressing to LV systolic impairment in a subset of patients in the presence of chronic volume and pressure overload.^34,35^ The lack of association with outcomes in the other LVH phenotypes is likely due to the relatively short follow-up period in UKB and relatively small population size of the test set. However, with the UKB aiming to scan 100,000 participants, there is potential to review outcomes in the future with greater numbers.

Nauta *et al* (2020) showed that patients with heart failure with eccentric LVH have a clinical and biomarker phenotype that is distinctly different from those with concentric LVH.^36^ In a retrospective post-hoc analysis of 1,015 patients with heart failure (LV ejection fraction <40%), the majority of patients (N=873) had eccentric LVH and were, on average, younger (P=0.005) and had a lower ejection fraction (P<0.001). The authors also found that beta-blocker up-titration was associated with a mortality benefit in heart failure with eccentric but not concentric LVH (P<0.001). Hypertension is an important risk factor for heart failure, therefore, improvements in screening are essential to reduce its burden and associated morbidity.^37^

Although the results are promising, further development and testing are required before implementation. For clinical applicability, the ML methods would need to be integrated into a point-of-care application or directly into ECG machines. Developing a model for single lead ECG would also be of interest, particularly in the era of wearable and smartphone technology. Considering the high global burden of hypertension, a cost-effective and accurate risk prediction of LVH may facilitate population screening and timely treatment in individuals with subclinical disease and could serve as surrogate markers for predicting outcomes. We have shown that eccentric LVH classified by ML is strongly associated with heart failure. This provides an opportunity to enhance targeted and personalized therapy for improvement in clinical outcomes of heart failure.

.The rate of new blood pressure-lowering therapies reaching the market has substantially declined, in part due to a lack of new targets for investigation. There is current interest in exploring the anti-fibrotic potential of sacubitril/valsartan (an angiotensin receptor-neprilysin inhibitor) in a clinical population, guided by imaging. REVERSE-LVH is a prospective, randomised, blinded endpoint (PROBE) clinical trial, designed to compare the effects of 52 weeks of treatment with sacubitril/valsartan with valsartan (an angiotensin receptor inhibitor) on the primary endpoint of change in interstitial volume measured by CMR in patients with hypertension and LVH.^38^ The UKB repeat imaging study may be an avenue to explore ECG-based classifiers in the reversal of LVH in hypertension.

Over thirty percent of adults worldwide have hypertension. Therefore, the potential cost-benefit of early detection of hypertension-mediated LVH is immense. Hypertension guidelines recommend the 12-lead ECG is performed in all patients newly diagnosed with hypertension.^39^ This provides an opportunity to compare the cost-benefit of ECG-based ML classification of LVH to that of conventional management. Important challenges will be to compare different sensitivity and specificity thresholds for the model to balance the trade-off between diagnostic accuracy and the economic benefit of downstream testing with potential false positive results. In order to implement an ECG-based ML screening strategy for LVH, it will be important to evaluate the cost-effectiveness under various clinical and cost scenarios.

Although we were able to perform external validation, an important limitation is that both cohorts are predominantly White European ancestry, therefore, further work is warranted to elucidate the classification of hypertension-mediated LVH in other ethnicities. Furthermore, our experiments included only ECG and clinical characteristics as features in the ML models. The rationale for this was to incorporate features that are accessible in a wide range of healthcare settings. Nevertheless, there is potential to include additional features to improve model performance and further personalize the ML algorithm. The UKB has access to numerous biomarkers and healthcare data such as BP medication history, biochemistry results, metabolomics, and genetic data, including genetic risk scores. These data could be incorporated into models for further development. In this study, we used three supervised ML approaches, and these models can be developed further to increase accuracy. Models using XGBoost (Extreme Gradient Boosting), decision trees and K-nearest neighbor are attractive options.^40–42^ Agnostic approaches such as unsupervised ML and deep learning (DL) may also be used, and these may identify novel signals in the ECG associated with LVH.

## Perspectives

This study demonstrates the potential of ECG-predicted ML classifiers to detect hypertension-mediated LVH phenotypes. A ML tool to detect hypertension-mediated LVH phenotypes may enhance clinician ECG interpretation and expedite workflow by ensuring that advanced imaging tests are used for those who need it most, thereby reducing unnecessary testing and subsequent waiting times. ML models based on ECG predictors offer new opportunities for improved and potentially cost-effective LVH detection, enhancing the capabilities of non-specialists. Future work will require validation testing in ethnically diverse cohorts with efforts to continue to improve performance metrics using auxiliary tools and health economics studies to assess the cost-benefit of this approach.

## Conclusions

ECG-based classifiers could discriminate between the four hypertension-mediated LVH phenotypes with external validation demonstrating robustness. We also observed a strong association between the ECG-predicted eccentric LVH and heart failure indicating there is important prognostic information gained from the model. This automated approach enhances the capabilities of non-specialists and potentially represents an accessible screening strategy for the early detection of hypertensives with LVH.

## Novelty and Relevance

### What is new?

- Supervised ML techniques using ECG and clinical data can detect LVH and discriminate between hypertension-mediated LVH phenotypes.
- The ECG provides important prognostic information and may provide clinicians with valuable information to assess CVD risk in hypertensives, particularly in low-resource settings.
- This automated approach may represent an efficient and accessible screening strategy for detecting subclinical LVH in hypertensives.

### What is relevant?

- Hypertension clinical guidelines endorse using the 12-lead ECG in hypertensives to screen for LVH.
- The diagnosis of hypertension-mediated LVH has relied on cardiac imaging, such as echocardiography and CMR.
- The potential cost implications of deploying and interpreting these imaging modalities as a screening strategy to detect hypertension-mediated LVH is not economically feasible.

## Clinical/Pathophysiological implications?

The ECG is a ubiquitous and low-cost diagnostic tool. In recent years, the transition to digitized ECG in electronic healthcare records and wearable technology has paved opportunities for ECG-based diagnostic and prognostic predictions. Early detection of hypertension-mediated LVH can enable regular healthcare follow-up, rigorous CV risk management and timely initiation of effective BP-reducing therapies.

## Ethics statement

This study complies with the Declaration of Helsinki; the work was covered by the ethical approval for UK Biobank studies from the NHS National Research Ethics Service on 17th June 2011 (Ref 11/NW/0382) and extended on 18 June 2021 (Ref 21/NW/0157) with written informed consent obtained from all participants. The work related to Study of Health in Pomerania is via application reference number SHIP/2023/31/D. The study is covered by the overall ethical approval for SHIP studies approved by the Ethics Committee at the University Medicine Greifswald, Germany.

## Non-standard Abbreviations and Acronyms

AI: Artificial intelligence
AUC: Area under the curve
BP: Blood pressure
CHD: Coronary heart disease
CI: Confidence interval
CMR: Cardiac magnetic resonance imaging
CV: Cardiovascular
CVD: Cardiovascular disease
DBP: Diastolic blood pressure
ECG: Electrocardiogram
HCM: Hypertrophic cardiomyopathy
LR: Logistic Regression
LV: Left ventricle
LVH: Left ventricular hypertrophy
MICE: Multivariate Imputation by Chained Equations
ML: Machine learning
mmHg: Millimetre of mercury
MRI: Magnetic resonance imaging
P: P-value
RF: Random forest
ROC: Receiver operator curves
SBP: Systolic blood pressure
SHIP: Study of Health in Pomerania
SVM: Support vector machine
UKB: UK Biobank

## Data Availability Statement

The data underlying this article were provided by the UK Biobank under access application 2964. UK Biobank will make the data available to bona fide researchers for all types of health-related research that is in the public interest, without preferential or exclusive access for any persons. All researchers will be subject to the same application process and approval criteria as specified by UK Biobank. For more details on the access procedure, see the UK Biobank website: http://www.ukbiobank.ac.uk/register-apply/.

## Acknowledgements

This study was conducted using the UK Biobank resource under access application 2964. We would like to thank all the participants, staff involved with planning, collection and analysis, including core lab analysis of the CMR imaging data. Graphical abstract was created using Biorender.com.

## Supplementary material

**Supplementary Table 1.** UK Biobank data fields used to identify hypertensive participants.

| Phenotype | Data fields | Field names | Data code definitions |
| --- | --- | --- | --- |
| Hypertension | 20002 | Non-cancer illness code, self-reported | Hypertension, Essential hypertension |
| Hypertension | 6150 | Vascular/heart problems diagnosed by doctor | High blood pressure |
| Hypertension | 6177 | Medication for cholesterol, blood pressure or diabetes | Blood pressure medication |
| Hypertension | 6153 | Medication for cholesterol, blood pressure, diabetes or take exogenous hormones | Blood pressure medication |
| Hypertension | 4080 | Systolic blood pressure, automated reading | $\geq 130$ mmHg |
| Hypertension | 4079 | Diastolic blood pressure, automated reading | $\geq 85$ mmHg |
| Hypertension | 93 | Systolic blood pressure, manual reading | $\geq 130$ mmHg |
| Hypertension | 94 | Diastolic blood pressure, manual reading | $\geq 85$ mmHg |
| Aortic stenosis | 20002 | Non-cancer illness code, self-reported | Aortic stenosis |
| Hypertrophic cardiomyopathy | 20002 | Non-cancer illness code, self-reported | Hypertrophic cardiomyopathy |

**Supplementary Table 2.** Definition of ECG biomarkers associated with LVH.

| ECG marker | Definition |
| --- | --- |
| Sokolow-Lyon index | (SV1 or SV2) + (RV5 or RV6) >35mm or R wave in aVL ≥11mm |
| Cornell voltage | SV3 +RaVL >28mm (men) >20mm (women) |
| Pathological Q waves | >30ms in duration and >1/3 of the R wave in depth in two or more contiguous leads (I, II, V1-6) |
| R wave amplitude (mV) | Amplitude of R wave in V5 or V6 >2.6mV, in aVL>1.1mV |
| QRS amplitude (mV) | Absolute value of the difference between maximum and minimum of the QRS complex values |
| QRS duration (ms) | QRS width from beginning of Q wave to end of S wave, (≥90ms) |
| QRS ascending slope | Upward slope of QRS complex |
| QRS descending slope | Downward slope of QRS complex |
| QTc interval (ms) | Prolonged corrected QT ≥450ms (men) ≥460ms (women) |
| STT segment amplitude | Absolute value of the difference between maximum and minimum of the ST segment values |
| ST segment displacement | Difference between the beginning of the QRS complex and the beginning of the T wave |
| QTc dispersion | Inter lead variations in QT segment length (<40ms) |
| T wave inversion | T wave amplitude $\leq -0.1$mV or a negative inflexion of at least 0.1mV in lead I, II or V3-6 |
| T wave amplitude (mV) | Absolute value of the difference between maximum and minimum of the T wave complex values |
| T wave axis ($^{\circ}$) | $<-15^{\circ}$ to $\geq-180^{\circ}$ or $>105^{\circ}$ to $\leq 180^{\circ}$ |
| T peak to T end interval (ms) | Duration between the peak of the T wave and the end of the T wave in each lead |

**Supplementary Table 3.** Codes used to define clinical outcomes using ICD-9, ICD-10and OPCS4 codes.

| **Myocardial Infarction** |  |
| --- | --- |
| **ICD10 codes** | **Definition** |
| I21 | Acute myocardial infarction |
| I21.0 | Acute transmural myocardial infarction of anterior wall |
| I21.1 | Acute transmural myocardial infarction of inferior wall |
| I21.2 | Acute transmural myocardial infarction of other sites |
| I21.3 | Acute transmural myocardial infarction of unspecified site |
| I21.4 | Acute subendocardial myocardial infarction |
| I21.9 | Acute myocardial infarction, unspecified |
| I22 | Subsequent myocardial infarction |
| I22.0 | Subsequent myocardial infarction of anterior wall |
| I22.1 | Subsequent myocardial infarction of inferior wall |
| I22.8 | Subsequent myocardial infarction of other sites |
| I22.9 | Subsequent myocardial infarction of unspecified site |
| I23 | Certain current complications following acute myocardial infarction |
| I23.0 | Haemopericardium as current complication following acute myocardial infarction |
| I23.1 | Atrial septal defect as current complication following acute myocardial infarction |
| I23.2 | Ventricular septal defect as current complication following acute myocardial infarction |
| I23.3 | Rupture of cardiac wall without haemopericardium as current complication following acute myocardial infarction |
| I23.5 | Rupture of papillary muscle as current complication following acute myocardial infarction |
| I23.6 | Thrombosis of atrium, auricular appendage and ventricle as current complications following acute myocardial infarction |
| I23.8 | Other current complications following acute myocardial infarction |
| I24 | Other acute ischaemic heart disease |
| I24.0 | Coronary thrombosis not resulting in myocardial infarction |
| I24.1 | Dressler's syndrome |
| I24.8 | Other forms of acute ischaemic heart disease |
| I24.9 | Acute ischaemic heart disease, unspecified |
| I25 | Chronic ischaemic heart disease |
| I25.0 | Atherosclerotic cardiovascular disease |
| I25.1 | Atherosclerotic heart disease |
| I25.2 | Old myocardial infarction |
| I25.3 | Aneurysm of heart |
| I25.4 | Coronary artery aneurysm |
| I25.5 | Ischaemic cardiomyopathy |
| I25.6 | Silent myocardial ischaemia |
| I25.8 | Other forms of chronic ischaemic heart disease |
| I25.9 | Chronic ischaemic heart disease, unspecified |
| **ICD9 codes** | **Definition** |
| 4109 | Acute myocardial infarction |
| **OPCS4** | **Definition** |
| K40 | Saphenous vein graft replacement of coronary artery |
| K40.1 | Saphenous vein graft replacement of one coronary artery |
| K40.2 | Saphenous vein graft replacement of two coronary arteries |
| K40.3 | Saphenous vein graft replacement of three coronary arteries |
| K40.4 | Saphenous vein graft replacement of four or more coronary arteries |
| K40.8 | Other specified saphenous vein graft replacement of coronary artery |
| K40.9 | Unspecified saphenous vein graft replacement of coronary artery |
| K41 | Other autograft replacement of coronary artery |
| K41.1 | Autograft replacement of one coronary artery NEC |
| K41.2 | Autograft replacement of two coronary arteries NEC |
| K41.3 | Autograft replacement of three coronary arteries NEC |
| K41.4 | Autograft replacement of four or more coronary arteries NEC |
| K41.8 | Other specified other autograft replacement of coronary artery |
| K41.9 | Unspecified other autograft replacement of coronary artery |
| K42 | Allograft replacement of coronary artery |
| K42.1 | Allograft replacement of one coronary artery |
| K42.2 | Allograft replacement of two coronary arteries |
| K42.3 | Allograft replacement of three coronary arteries |
| K42.4 | Allograft replacement of four or more coronary arteries |
| K42.8 | Other specified other allograft replacement of coronary artery |
| K42.9 | Unspecified other allograft replacement of coronary artery |
| K44 | Other replacement of coronary artery |
| K44.1 | Replacement of coronary arteries using multiple methods |
| K44.2 | Revision of replacement of coronary artery |
| K44.8 | Other specified other replacement of coronary artery |
| K44.9 | Unspecified other replacement of coronary artery |
| K45 | Connection of thoracic artery to coronary artery |
| K45.1 | Double anastomosis of mammary arteries to coronary arteries |
| K45.2 | Double anastomosis of thoracic arteries to coronary arteries NEC |
| K45.3 | Anastomosis of mammary artery to left anterior descending coronary artery |
| K45.4 | Anastomosis of mammary artery to coronary artery NEC |
| K45.5 | Anastomosis of thoracic artery to coronary artery NEC |
| K45.6 | Revision of connection of thoracic artery to coronary artery |
| K45.8 | Other specified connection of thoracic artery to coronary artery |
| K45.9 | Unspecified connection of thoracic artery to coronary artery |
| K49 | Transluminal balloon angioplasty of coronary artery |
| K49.1 | Percutaneous transluminal balloon angioplasty of one coronary artery |
| K49.2 | Percutaneous transluminal balloon angioplasty of multiple coronary arteries |
| K49.3 | Percutaneous transluminal balloon angioplasty of bypass graft of coronary artery |
| K49.4 | Percutaneous transluminal cutting balloon angioplasty of coronary artery |
| K49.8 | Other specified transluminal balloon angioplasty of coronary artery |
| K49.9 | Unspecified transluminal balloon angioplasty of coronary artery |
| K50 | Other therapeutic transluminal operations on coronary artery |
| K50.1 | Percutaneous transluminal laser coronary angioplasty |
| K50.2 | Percutaneous transluminal coronary thrombolysis using streptokinase |
| K50.3 | Percutaneous transluminal injection of therapeutic substance into coronary artery NEC |
| K50.4 | Percutaneous transluminal atherectomy of coronary artery |
| K50.8 | Other specified other therapeutic transluminal; operations on coronary artery |
| K50.9 | Unspecified other therapeutic transluminal operations on coronary artery |
| K75 | Percutaneous transluminal balloon angioplasty and insertion of stent into coronary artery |
| K75.1 | Percutaneous transluminal balloon angioplasty and insertion of 1-2 drug-eluting stents into coronary artery |
| K75.2 | Percutaneous transluminal balloon angioplasty and insertion of 3 or more drug-eluting stents into coronary artery |
| K75.3 | Percutaneous transluminal balloon angioplasty and insertion of 1-2 stents into coronary artery |
| K75.4 | Percutaneous transluminal balloon angioplasty and insertion of 3 or more stents into coronary artery NEC |
| K75.8 | Other specified percutaneous transluminal balloon angioplasty and insertion of stent into coronary artery |
| K75.9 | Unspecified percutaneous transluminal balloon angioplasty and insertion of stent into coronary artery |
| **Heart Failure** |  |
| **ICD10 codes** | **Definition** |
| I11.0 | Hypertensive heart disease with (congestive) heart failure |
| I13.0 | Hypertensive heart and renal disease with (congestive) heart failure |
| I13.2 | Hypertensive heart and renal disease with both (congestive) heart failure and renal failure |
| I25.5 | Ischaemic cardiomyopathy |
| I50 | Heart failure |
| I50.0 | Congestive heart failure |
| I50.1 | Left ventricular failure |
| I50.9 | Heart failure, unspecified |
| J81 | Pulmonary oedema |
| K76.1 | Chronic passive congestion of liver |
| **ICD9 codes** | **Definition** |
| 4280 | Congestive heart failure |
| 4281 | Left heart failure |
| 4289 | Heart failure, unspecified |
| **OPCS4 codes** | **Definition** |
| K59.6 | Implantation of cardioverter defibrillator using three electrode leads |
| K61.7 | Implantation of biventricular cardiac pacemaker system |
| K60.7 | Implantation of intravenous biventricular cardiac pacemaker system |
| **Stroke** |  |
| **ICD10 codes** | **Definition** |
| I60 | Subarachnoid haemorrhage |
| I60.0 | Subarachnoid haemorrhage from carotid siphon bifurcation |
| I60.1 | Subarachnoid haemorrhage from middle cerebral artery |
| I60.2 | Subarachnoid haemorrhage from anterior communicating artery |
| I60.3 | Subarachnoid haemorrhage from posterior communicating artery |
| I60.4 | Subarachnoid haemorrhage from basilar artery |
| I60.5 | Subarachnoid haemorrhage from vertebral artery |
| I60.6 | Subarachnoid haemorrhage from other intracranial arteries |
| I60.7 | Subarachnoid haemorrhage from intracranial artery, unspecified |
| I60.8 | Other subarachnoid haemorrhage |
| I60.9 | Subarachnoid haemorrhage, unspecified |
| I61 | Intracerebral haemorrhage |
| I61.0 | Intracerebral haemorrhage in hemisphere subcortical |
| I61.1 | Intracerebral haemorrhage in hemisphere, cortical |
| I61.2 | Intracerebral haemorrhage in hemisphere, unspecified |
| I61.3 | Intracerebral haemorrhage in brain stem |
| I61.4 | Intracerebral haemorrhage in cerebellum |
| I61.5 | Intracerebral haemorrhage, intraventricular |
| I61.6 | Intracerebral haemorrhage, multiple localised |
| I61.8 | Other intracerebral haemorrhage |
| I61.9 | Intracerebral haemorrhage, unspecified |
| I62 | Other nontraumatic intracranial haemorrhage |
| I62.0 | Subdural haemorrhage (acute) (nontraumatic) |
| I62.1 | Nontraumatic extradural haemorrhage |
| I62.9 | Intracranial haemorrhage (nontraumatic), unspecified |
| I63 | Cerebral infarction |
| I63.0 | Cerebral infarction due to thrombosis of precerebral arteries |
| I63.1 | Cerebral infarction due to embolism of precerebral arteries |
| I63.2 | Cerebral infarction due to unspecified occlusion or stenosis of precerebral arteries |
| I63.3 | Cerebral infarction due to thrombosis of cerebral arteries |
| I63.4 | Cerebral infarction due to embolism of cerebral arteries |
| I63.5 | Cerebral infarction due to unspecified occlusion or stenosis of cerebral arteries |
| I63.6 | Cerebral infarction due to cerebral venous thrombosis, nonpyogenic |
| I63.8 | Other cerebral infarction |
| I63.9 | Cerebral infarction, unspecified |
| I64 | Stroke, not specified as haemorrhage or infarction |
| I65 | Occlusion and stenosis of precerebral arteries, not resulting in cerebral infarction |
| I65.0 | Occlusion and stenosis of vertebral artery |
| I65.1 | Occlusion and stenosis of basilar artery |
| I65.2 | Occlusion and stenosis of carotid artery |
| I65.3 | Occlusion and stenosis of multiple and bilateral precerebral arteries |
| I65.8 | Occlusion and stenosis of other precerebral artery |
| I65.9 | Occlusion and stenosis of unspecified precerebral artery |
| I66 | Occlusion and stenosis of cerebral arteries, not resulting in cerebral infarction |
| I66.0 | Occlusion and stenosis of middle cerebral artery |
| I66.1 | Occlusion and stenosis of anterior cerebral artery |
| I66.2 | Occlusion and stenosis of posterior cerebral artery |
| I66.3 | Occlusion and stenosis of cerebellar arteries |
| I66.4 | Occlusion and stenosis of multiple and bilateral cerebral arteries |
| I66.8 | Occlusion and stenosis of other cerebral artery |
| I66.9 | Occlusion and stenosis of unspecified cerebral artery |
| I67.0 | Dissection of cerebral arteries, nonruptured |
| I67.8 | Other specified cerebrovascular diseases |
| I67.9 | Cerebrovascular disease, unspecified |
| I69 | Sequelae of cerebrovascular disease |

| ICD9 codes | Definition |
| --- | --- |
| 4309 | Subarachnoid haemorrhage |
| 4319 | Intracerebral haemorrhage |
| 4320 | Nontraumatic extradural haemorrhage |
| 4321 | Subdural haemorrhage |
| 4331 | Occlusion and stenosis of carotid artery |
| 4339 | Occlusion and stenosis of precerebral arteries, unspecified |
| 4349 | Occlusion of cerebral arteries, unspecified |
| 4369 | Acute but ill-defined cerebrovascular disease |
| 4371 | Other generalised ischaemic cerebrovascular disease |
| OPCS4 codes | Definition |
| L35.4 | Percutaneous transluminal embolectomy of cerebral artery |
| Ventricular Arrhythmias |  |
| ICD10 codes | Definition |
| I47.2 | Ventricular tachycardia |
| I49.0 | Ventricular fibrillation and flutter |
| I46.0 | Cardiac arrest with successful resuscitation |
| I46.1 | Sudden cardiac death, so described |
| I46.9 | Cardiac arrest, unspecified |
| I47.0 | Re-entry ventricular arrhythmia |
| ICD9 codes | Definition |
| 4270 | Paroxysmal ventricular tachycardia |
| 4271 | Paroxysmal ventricular tachycardia |
| 4272 | Paroxysmal tachycardia, unspecified |
| 4274 | Ventricular fibrillation and flutter |
| OPCS4 codes | Definition |
| K59 | Cardioverter defibrillator introduced through vein |
| K59.1 | Implantation of cardioverter defibrillator using one electrode lead |
| K59.2 | Implantation of cardioverter defibrillator using two electrode leads |
| K59.3 | Resitting of leads of cardioverter defibrillator |
| K59.4 | Renewal of cardioverter defibrillator |
| K59.6 | Implantation of cardioverter defibrillator using three electrode leads |
| K59.8 | Other specified cardioverter defibrillator introduced through the vein |
| K59.9 | Unspecified cardioverter defibrillator introduced through the vein |
| K72 | Other cardioverter defibrillator |
| K72.1 | Implantation of subcutaneous cardioverter defibrillator |
| K72.3 | Renewal of subcutaneous cardioverter defibrillator |

**Supplementary Table 4.** Associations of ECG-predicted hypertension-mediated LVH phenotypes and clinical outcomes.

|  | **MACE** |  | **Heart Failure** |  |
| --- | --- | --- | --- | --- |
|  | HR (95% CI) | P-value | HR (95% CI) | P-value |
| **LV remodeling** | 1.40 (0.37-2.76) | 0.1 | 1.55 (0.71-3.36) | 0.3 |
| **Eccentric LVH** | 0.93 (0.42-2.04) | 0.9 | 3.24 (1.06-9.86) | 0.04 |
| **Concentric LVH** | 1.01 (0.37-2.76) | 0.1 | 1.27 (0.17-9.65) | 0.8 |

*AUC: area under the receiver operator curve; CI: confidence interval; HR: hazard ratio; MACE: major adverse cardiovascular events; LVH: left ventricular hypertrophy.*

**Supplementary figure 1.**
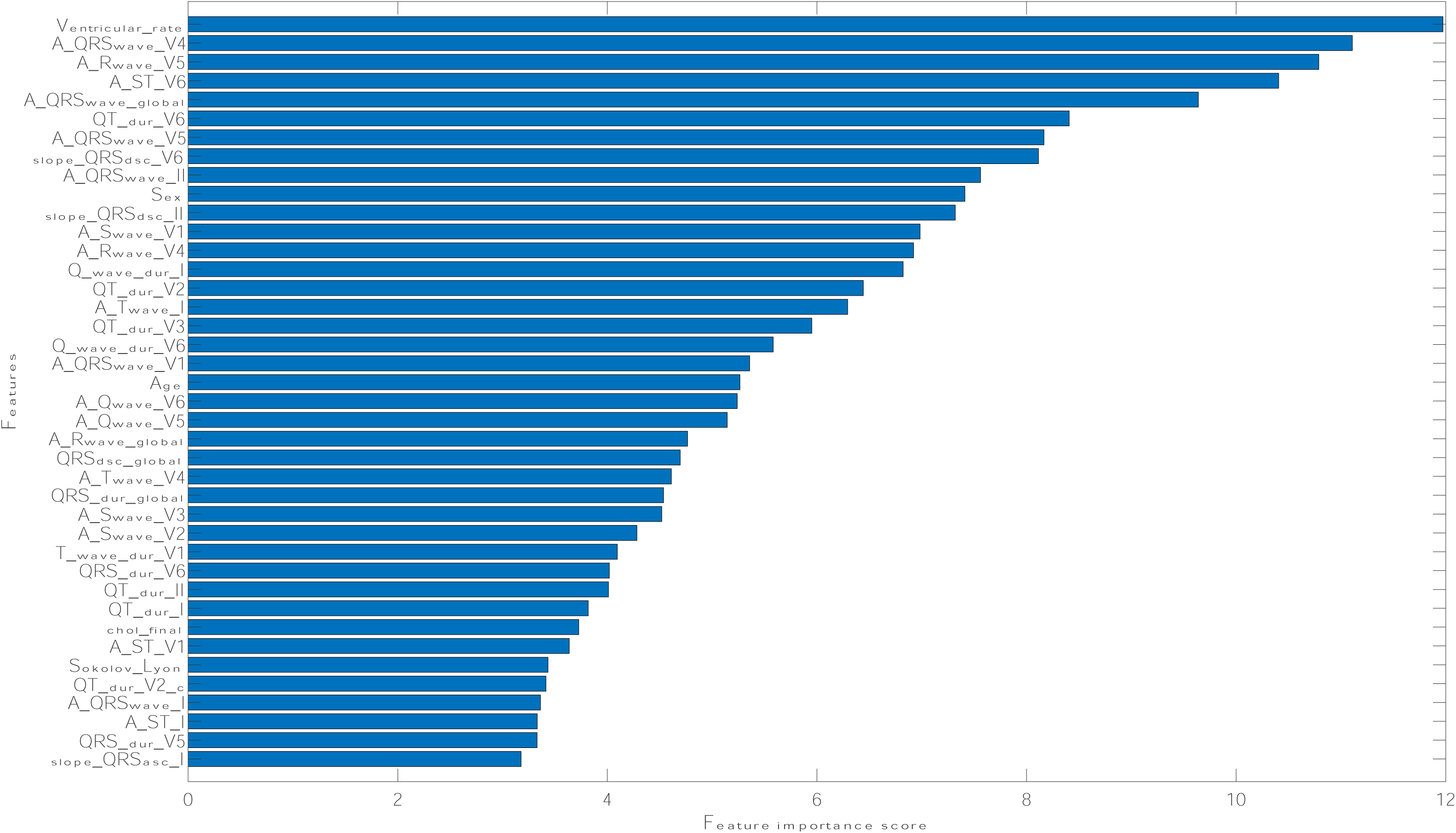
Ranking of the top 40 features using chi-squared feature selection.

